# Identification of the Risk Factors for Complex-Multiple Long-term Conditions (C-MLTC): A Scoping Review

**DOI:** 10.1101/2024.07.08.24310078

**Authors:** Carl J. Heneghan, Jon Brassey, James P. Sheppard, Apostolos Tsiachristas, Kamaldeep Bhui, Rafael Perera, Derrick Bennett, Igho J. Onakpoya

## Abstract

**Background:** Complex multiple long-term conditions (C-MLTC) are commonly defined as the presence of four or more long-term conditions, and their prevalence is projected to increase significantly over the next decade.

**Methods:** We undertook a scoping review of the literature to identify available evidence on the risk factors for C-MLTC and map out the potential clusters. We focussed on a subset of 18 chronic conditions because they include the most commonly reported and prevalent conditions in the general population. For guidelines we searched NICE Clinical Knowledge Summaries (CKS) and for systematic reviews we searched PubMed, Google Scholar and the TRIP database.

**Results:** We found 53 risk factors associated with 18 long-term conditions. Sixteen of these are linked to C-MLTC, with 11 being modifiable. Although we reviewed 51 systematic reviews, inconsistent reporting made it difficult to quantify the risk factors and conditions associated with C-MLTC. Diabetes mellitus was a risk factor for most other conditions (10), followed by coronary heart disease, hypertension, and chronic kidney disease (each associated with 8 long-term conditions). Additionally, depression and anxiety are commonly associated with other long-term conditions, with 19 and 20 conditions, respectively.

**Conclusion:** Several risks and conditions contribute to the development of multimorbidity. Anxiety and depression are nearly associated with all long-term conditions. Reporting problems within the existing evidence and a lack of clear definitions prevent adequate quantification of the risks related to C-MLTC.

## INTRODUCTION

One in four adults in England has Multiple Long-Term Conditions (MLTC), which is the co-occurrence of two or more diseases in the same individual, each of which must be a non-communicable disease (NCD) [1]. NCDs account for roughly 70% of all deaths globally [2] and are more common with ageing, affecting two-thirds of the elderly [3].

Multimorbidity is commonly understood to be the coexistence of multiple long-term conditions in an individual. However, its study is limited because the predominant research approach has been to study single diseases or through the lens of comorbidity. The concept of single disease characterisation dates back to Thomas Sydenham (Diseases 1624-89), who differentiated “Acute diseases meaning those of which God is the author, chronic meaning those that originate in ourselves” [4]. In 1957, the concept of “risk factors’’ in coronary heart disease (CHD) was first coined by the Framingham Heart Study (FHS). In 1970, Feinstein coined comorbidity [5], providing definitions that indicate two or more medical conditions existing simultaneously regardless of their causal relationship. Complex-MLTC (C-MLTC) is commonly defined as the presence of four or more long-term conditions; this is projected to increase to 17% of the adult population by 2035, partly due to the ageing population. The evidence shows that C-MLTC decreases the quality of life and increases morbidity and mortality [6,7].

Current management tends to focus on treating individual conditions, and single-disease guidelines often do not consider patients with multimorbidity [8]. Furthermore, the transition from single conditions to C-MLTC is poorly understood. For example, there is a lack of understanding of what increases the risk or speeds up the transition to C-MLTC, and there is limited evidence on the life course for progression to C-MLTC and whether specific interventions can decrease the risk [9]. There is a lack of evidence about whether overdiagnosis and polypharmacy increase the risk of or progression to C-MLTC, and there are uncertainties about the effectiveness of interventions generally due to a paucity of randomised controlled trials [10].

Complex-MLTC may occur due to different combinations of chronic conditions, and it is unclear how the health and economic burden to patients, their carers, and care systems is distributed across the various clusters of disease that may occur [11]. Research is emerging to identify which conditions cluster together, how the burden of C-MLTC relates to health and ageing, what risk factors influence the development of C-MLTC, and which chronic conditions affect the burden of disease. For example, a recent retrospective longitudinal (panel) study identified multimorbidity clusters with the highest primary care demand by consultation type and ethnicity [12].

We undertook a scoping review of the literature to identify the available evidence on the risk factors for C-MLTC and to map out potential clusters of C-MLTC. The objectives of the review were:

1. To identify the risk factors and conditions associated with C-MLTC.
2. To identify studies that report on the aetiology, development, and risk factors for C-MLTC.

## METHODS

The review protocol is available here: https://osf.io/vte3a/; and a previous pre-print is available here: https://www.medrxiv.org/content/10.1101/2024.07.08.24310078v1.

### Inclusion criteria

We focussed the review and outputs on a subset of 18 chronic conditions affecting the cardiovascular, renal, musculoskeletal and respiratory systems, mental health conditions and cancer. The 18 conditions are Stroke/TIA, Coronary heart disease (CHD), Atrial Fibrillation (AF), Heart Failure (HF), Hypertension, Peripheral Arterial Disease (PAD), Diabetes Mellitus Asthma, Chronic Obstructive Pulmonary Disease (COPD), Dementia, Parkinson’s Disease; Depression; Anxiety; Serious Mental Illnesses (Bipolar Disorder and Schizophrenia); Cancer excluding non-melanoma skin cancers; Chronic Kidney Disease (CKD); Osteoporosis; and Rheumatoid Arthritis (RA).

We focussed on this subset of 18 chronic conditions because they include the most commonly reported and prevalent in the general population, in mid-life (45-64 years) and in early old age (64-85 years) [13,14,15]. To achieve objective 1, we identified risk factors and conditions associated with C-MLTC; we cross-tabulated LTC risk factors by the individual condition based on NICE Clinical Knowledge Summaries (CKS) (https://cks.nice.org.uk/), tabulating the risk factors for each of the 18 conditions and the relationship of the 18 conditions as risk factors.

To achieve objective 2, we identified studies that reported on the aetiology, development and risk factors for C-MLTC and generated the following PECO criteria:

**P:** Individuals with one of the 18 long-term conditions

**E:** Risk factors (identified in Question 1)

**C:** No risk factor

**O:** Increase in the risk of C-MLTC

We then searched for systematic reviews that included MLTC and reported as an outcome of the development of MLTC across the 18 conditions that fulfilled the PECO criteria above. In addition, for a study to be eligible, study participants had to be 18 years or older, and the modifiable risk factor must have predated the outcome of one of the 18 specified diseases in adulthood. We included systematic reviews containing studies meeting our eligibility criteria. Where applicable, we included guidelines or guideline reports from the grey literature and only included studies published in English.

### Search strategy

We used NICE Clinical Knowledge Summaries (CKS) guidelines and for systematic reviews we searched the following electronic databases: PubMed, Google Scholar and Trip database. The search was conducted in each database from inception until 31 May 2023 and then updated to 31 October 2024. The search terms were developed iteratively using the 18 conditions as the starting point of their search. Search terms used included the individual conditions alongside the qualifiers *comorbidity* and *risk factors* and, where supported, appropriate MeSH terms. We also screened the reference lists of included studies for possible additional studies to assess eligibility and conducted citation searching.

### Selection of sources of evidence

Three reviewers (CJH, JB, and IJO) screened the initial studies retrieved for eligibility. The first screen was based on the study title and abstract; the next screen used the full text to assess inclusion. Any disagreements were resolved by consensus or discussion with a third reviewer if necessary. The inclusion and exclusion of retrieved studies were reported in a flow diagram according to PRISMA for scoping review reporting guidelines [16].

### Quality

We documented the quality assessment tool used in the reviews and reported the quality of the evidence as assessed in the original systematic reviews.

### Data extraction

We attempted to quantify the risk factors and conditions associated with C-MLTC and report the univariate analysis by conditions and, where reported, results adjusted by non-modifiable risk factors such as age, sex, family history and ethnicity. We extracted data into a template that consisted of the number of studies in the review, the number of participants, and the outcomes for the 18 conditions. One reviewer (IJO) extracted the data and checked with a second reviewer (CJH). Any disagreements were resolved via consensus.

### Synthesis and dissemination of results

The scoping review is reported according to the PRISMA statement on reporting scoping reviews [16], and the data is made available on the Open Science Framework [17].

## RESULTS

### Risk factors for C-MLTC

We found 53 risk factors in the NICE CKS guidance associated with at least one of the 18 Individual Long-Term Conditions (LTCs). Supplementary Table 1a shows that CKS reports 23 risk factors associated with multimorbidity (two or more LTCs). Of these, 16 risks are associated with C-MLTC (four or more LTCs), of which 11 are modifiable risk factors. Cigarette smoking was the modifiable risk factor associated with most LTCs (n=11), followed by obesity (n=9) and blood pressure (n=8). NICE CKS reports 30 risk factors associated with only one of the 18 LTCs. (see Supplementary Table 1b).

From our initial search of 1482 studies identified from the four different databases. Our updated searches identified an additional 33 studies, bringing the total to 950 non-duplicate studies (see Figure 1 flow diagram). We excluded 756 studies on title and abstract and retrieved 194 reports for full-text screening. Of these, we assessed 97 in detail, excluding 23 as they had no extractable information and five because, on further analysis, they were out of scope. Therefore, we included 51 SRs [18–68] with 1,789 primary studies (range 4 to 248, median 20 studies). Two reviews (Lanctôt 2023 [39]; Roy 2012 [58]) included both systematic reviews and primary studies, while one review [Brain 2023] was an overview of meta-analytic reviews. These studies included 136,888,649 participants across 42 studies (range 205 to 83,020,812). The total number of participants in nine SRs was unclear. Supplementary Table 2a reports the systematic reviews’ extractable information.

Forty reviews assessed the quality of the studies (see Supplementary Table 2a). Eighteen assessment tools were used, with the Newcastle Ottawa tool being the most frequent (n=16 studies). In general, most of the reviews reported the overall quality as moderate to high.

Three reviews (Huang 2020a [33]; Khaledi 2019 [38]; Pahwa 2023 [54]) did not specify the overall reporting quality of the primary studies, and the authors of one review [65] used an assessment tool that has not been validated. One review [28] excluded primary studies with low quality.

Overall, due to inconsistency within the included studies, the results did not allow us to quantify the risk factors and conditions associated with C-MLTC adequately. Twenty studies quantified the risk of the long-term conditions under study (Supplementary Table 2b). In the reviews we included, no pooled associations were reported for the risk of developing COPD, asthma, bipolar affective disorder, osteoporosis, or rheumatoid arthritis. Dementia was the most reported pooled outcome in seven reviews.

When quantifiable results were available, they illustrated the complex interplay long-term conditions have on each other’s development. For example, Huang 2020a showed that COPD patients with heart failure were at significant risk of new-onset AF, whereas other conditions were not.

### Long-term Conditions as Risk Factors for C-MLTC

Supplementary Table 3 (Long-term Conditions as Risk Factors for Multimorbidity) shows the conditions acting as risk factors for the other 17 conditions. In the table, we removed the association of a condition with itself to give the total number of long-term conditions to which the risk condition contributes. The results show that all but one condition (asthma) is a risk factor for multimorbidity, with ten conditions associated with C-MLTC. We found that diabetes mellitus is a risk factor for most other conditions (n=10), followed by coronary heart disease, hypertension, and chronic kidney disease (n=8 long-term conditions each). We also found that depression and anxiety are the most other long-term conditions as risk factors (14 and 17, respectively); they contribute little to the risk of developing other conditions.

Counter to the CKS results, Becker 2018 [21] reported that anxiety predicts the risk of increased risk of Alzheimer’s disease (HR 1.53, 95% CI 1.16-2.01) and vascular dementia (odds ratio 1.88, 95% CI 1.05-3.36, P < 0.01). Also, Kuring 2020 [38] reported a significantly higher risk of developing all-cause dementia (OR 1.91, 1.72-2.12) and AD (OR 1.60,1.29-2.00) in people with previous depression. In Hotzel 2011 [31], anxiety disorders and personality disorders appeared concurrently with chronic depression. In Huang’s 2020 review [32], CKD plus concomitant depressive symptoms (10 studies) or comorbidity index score (5 studies) increased the risk of anxiety. Khaledi’s 2019 review of 248 studies [37], including 83 million patients, reported a pooled prevalence of depression among patients with T2DM in the world of 28.0% (95% CI 27-29).

Furthermore, Naqvi 2019 [47] showed that 47% of rheumatoid arthritis patients had comorbidities. Undiagnosed depression was commonly reported in 58.3% of the patients. However, Steck 2018 [60] highlights that the impact of depression varies depending on the review and the studies included.

## DISCUSSION

We found 23 risk factors that contribute to the development of multimorbidity, and 16 are associated with C-MLTC, of which 11 are modifiable risk factors. We also showed that all but one condition (asthma) are risk factors for multimorbidity, and ten conditions are associated with MLTC. We found that diabetes mellitus is a risk factor for ten other conditions, followed by coronary heart disease, hypertension, and chronic kidney disease (n=8 long-term conditions each). Our results also show that nearly all long-term conditions act as risks for depression and anxiety. However, due to reporting problems, our results did not allow us to quantify the risk factors and conditions associated with C-MLTC, and depending on the studies chosen, results differed in magnitude and due to a lack of reporting of the temporality we cannot determine the direction of these associations.

Multimorbidity is a relatively new concept compared to single or co-morbid diseases. In 2018, the Lancet published Making More of Multimorbidity as an Emerging Priority [69], and the Academy of Medical Sciences [70] highlighted that multimorbidity was a priority for global health research. In 2016, NICE guidance on the clinical assessment and management of multimorbidity highlighted evidence for recommendations in NICE guidance on single health conditions, which are regularly drawn from people without multimorbidity and taking fewer prescribed regular medicines [71].

Consequently, numerous areas of contention make multimorbidity complex to study. There has yet to be an international consensus regarding defining and measuring multimorbidity. This makes conducting and interpreting research, comparing findings across populations, and developing guidelines and interventions complex. A review of prevalence studies of multimorbidity found estimates ranging from less than 5% to more than 95%, often due to differences in the operational definition of multimorbidity [72]. A cross-sectional study in Scotland extracted data on 40 morbidities and found that 42% of all patients had one or more morbidities, and 23% were multimorbid. While the prevalence of multimorbidity increased with age, the absolute number of people with multimorbidity was higher in those under 65 [73].

While multimorbidity is commonly understood to be the coexistence of multiple health conditions in an individual, and complex multiple long-term conditions (C-MLTC) is the coexistence of four or more health conditions, the exact components are ill-defined. For example, NICE has a more extensive list that constitutes multimorbidity conditions, including physical and mental health conditions such as diabetes or schizophrenia, ongoing conditions such as learning disability, symptom complexes such as frailty or chronic pain, sensory impairment such as sight or hearing loss, and alcohol and substance misuse. A systematic review emphasised the heterogeneity of existing multimorbidity indices. Of the 39 included studies in the review, more than half (59%) had no selection criteria for the conditions included. On average, the studies included 18.5 diseases, ranging between 4 and 102 different conditions [74], identifying more than 150 different conditions that might account for multimorbidity. However, several conditions, including diabetes mellitus, stroke, COPD, cancer, arthritis, and hypertension, were listed in nearly every study. The review concluded that an important similarity across studies is the focus on diseases with a high prevalence and a severe impact on affected individuals, as this present overview seeks to do.

### Limitations

Our approach has several limitations. First, we found it difficult to locate studies. The term multimorbidity has recently been defined, and the studies we chose to include often did not reflect what was included in many reviews. However, the conditions we focussed on include the most commonly reported and prevalent in the UK general population, in mid-life (45-64 years) and in early old age (64-85 years) [75,76,77]. Some studies have analysed the interaction of hundreds of conditions [78]. This approach is more comprehensive, but it remains to be seen if this process is generalisable and valuable for clinical practice. Furthermore, we found substantial problems with reporting and a lack of standard definitions and terminology. Inconsistent reporting occurs because most studies are set up to analyse comorbidities and not multimorbidity. It is also unclear whether systematic reviews are the way to study multimorbidity. We do not know the extent to which results can be generalised outside of the country setting by different characteristics such as ethnicity, differing disease definitions and ascertainment of cases. Furthermore, we were unable to assess the interaction of risk factors. For example, the odds of having peripheral arterial disease increase with higher numbers of risk factors. One risk factor gives a 1.5-fold increase, and three or more shows a 10-fold increase [79]. In addition, NICE CKS results are not backed up by a systematic approach to risk factor identification. They differ from the results of several systematic reviews in identifying risk factors.

### Implications for research

Our findings show the need for consistency in the definitions of long-term conditions and multimorbidity. An agreed glossary of definitions and reporting standards would aid the development of relevant evidence synthesis. A register of population-based studies on multimorbidity and reporting the methods used, the disease studied, and the outcomes of interest may prove a helpful resource. This is particularly important given the study of multimorbidity is in its infancy. There is a need to connect evidence from single diseases and comorbid conditions with the growing problem of multimorbidity and C-MLTC management. However, an attempt to synthesise the evidence on multimorbidity will be labour-intensive. For example, over ten thousand systematic reviews on diabetes mellitus are indexed in PubMed. The limitations in the generalisability of findings may mean that results may not transfer to other settings. Therefore, limiting results to country-specific settings may be more coherent when analysing the risks of multimorbidity and C-MLTC. In addition, universally accepted tools for assessing reporting quality of studies investigating C-MLTC should be developed to enable comparisons across studies.

## CONCLUSIONS

Our Scoping review shows that several risks and conditions contribute to the development of multimorbidity and C-MLTC. Anxiety and depression are nearly associated with all long-term conditions. Reporting problems within the systematic reviews and a lack of clear definitions prevented us from adequately quantifying the risk factors and conditions associated with C-MLTC.

## Data Availability

Data are made available online on the Open Science Framework

https://osf.io/vte3a/

## PRISMA 2020 flow diagram for new systematic reviews which included searches of databases and registers only

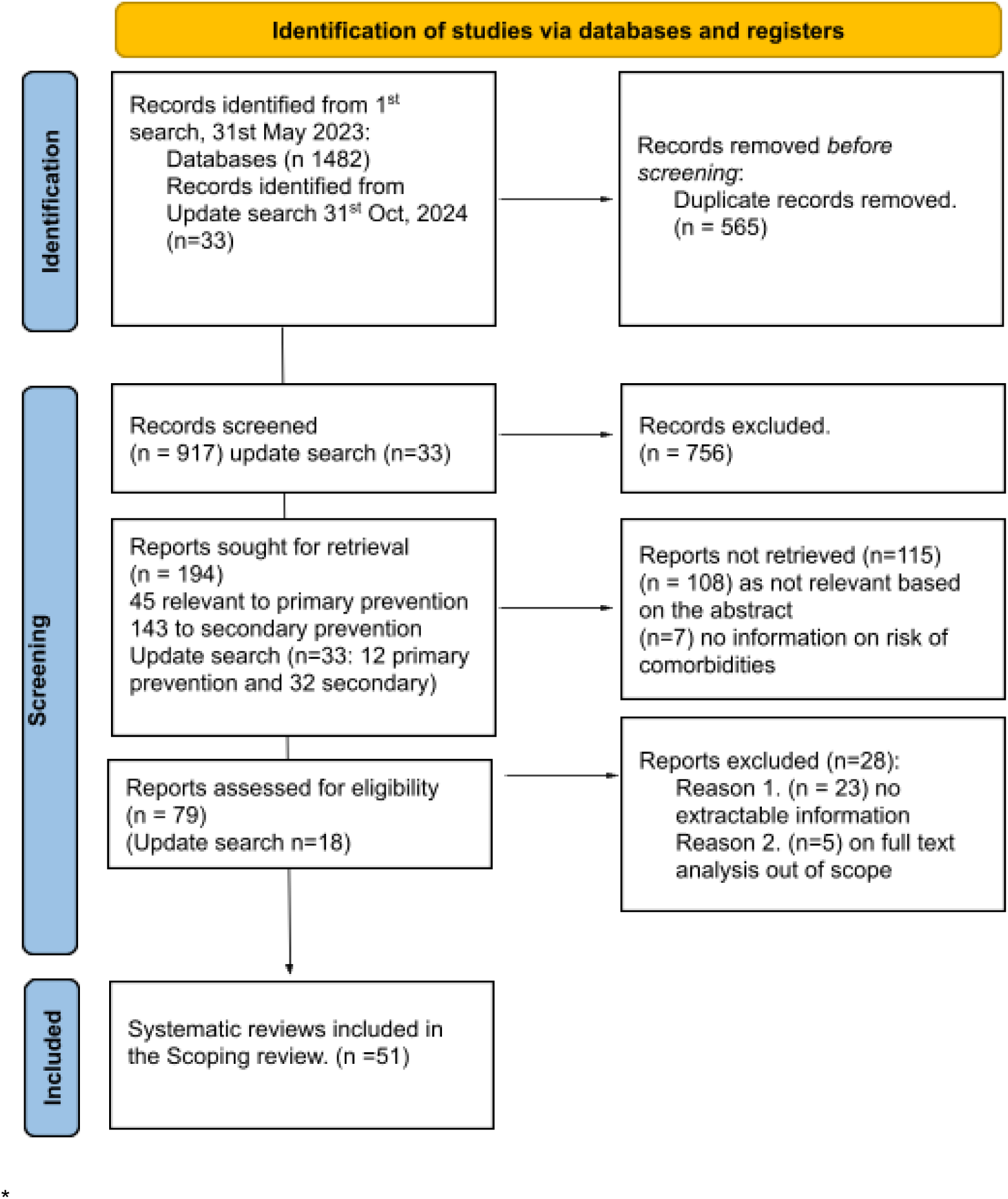

From: Page MJ, McKenzie JE, Bossuyt PM, Boutron I, Hoffmann TC, Mulrow CD, et al. The PRISMA 2020 statement: an updated guideline for reporting systematic reviews. BMJ 2021;372:n71. doi: 10.1136/bmj.n71 For more information, visit: http://www.prisma-statement.org/

