## Supplementary material for "Identification of the Risk Factors for Complex-Multiple Long-term Conditions (C-MLTC): A Scoping Review": Suppl. Table 1a

| **Suppl. Table 1a**  **Risk factors**  **for multimorbidity (n=23)** | Stroke/TIA | Coronary Heart disease | Atrial Fibrillation | Heart Failure | Hypertension | PAD | Diabetes Mellitus | Asthma | COPD | Dementia | Parkinson’s Disease | Depression | Anxiety | Bipolar Disorder | Schizophrenia | Prostate Cancer | Ovarian Cancer | Lung Cancer | Breast Cancer | Chronic Kidney Disease | Osteoporosis | Rheumatoid Arthritis | **Total** |
| --- | --- | --- | --- | --- | --- | --- | --- | --- | --- | --- | --- | --- | --- | --- | --- | --- | --- | --- | --- | --- | --- | --- | --- |
| **CKS Link** | [**Link**](https://cks.nice.org.uk/topics/stroke-tia/background-information/risk-factors/) | [**Link**](https://cks.nice.org.uk/topics/cvd-risk-assessment-management/background-information/risk-factors-for-cvd/) | [**Link**](https://cks.nice.org.uk/topics/atrial-fibrillation/background-information/causes/) | [**Link**](https://cks.nice.org.uk/topics/heart-failure-chronic/background-information/causes/) | [**Link**](https://cks.nice.org.uk/topics/hypertension/background-information/risk-factors/) | [**Link**](https://cks.nice.org.uk/topics/peripheral-arterial-disease/background-information/risk-factors/) | [**Link**](https://cks.nice.org.uk/topics/diabetes-type-2/background-information/risk-factors/) | [**Link**](https://cks.nice.org.uk/topics/asthma/background-information/risk-factors/) | [**Link**](https://cks.nice.org.uk/topics/chronic-obstructive-pulmonary-disease/background-information/risk-factors/) | [**Link**](https://cks.nice.org.uk/topics/dementia/background-information/risk-factors/) | [**Link**](https://cks.nice.org.uk/topics/parkinsons-disease/) | [**Link**](https://cks.nice.org.uk/topics/depression/background-information/risk-factors/) | [**Link**](https://cks.nice.org.uk/topics/generalized-anxiety-disorder/background-information/risk-factors/) | [**Link**](https://cks.nice.org.uk/topics/bipolar-disorder/background-information/causes/) | [**Link**](https://cks.nice.org.uk/topics/psychosis-schizophrenia/background-information/causes-risk-factors/) | [**Link**](https://cks.nice.org.uk/topics/prostate-cancer/background-information/risk-factors/) | [**Link**](https://cks.nice.org.uk/topics/ovarian-cancer/background-information/risk-factors/) | [**Link**](https://cks.nice.org.uk/topics/lung-pleural-cancers-recognition-referral/) | [**Link**](https://cks.nice.org.uk/topics/breast-cancer-managing-fh/management/breast-cancer-managing-fh/) | [**Link**](https://cks.nice.org.uk/topics/chronic-kidney-disease/background-information/causes/) | [**Link**](https://cks.nice.org.uk/topics/osteoporosis-prevention-of-fragility-fractures/background-information/risk-factors/) | [**link**](https://www.nice.org.uk/about/what-we-do/into-practice/measuring-the-use-of-nice-guidance/impact-of-our-guidance/nice-impact-arthritis/diagnosis-and-referral-of-inflammatory-arthritis) |  |
| Age | Yes | Yes |  |  | Yes | Yes |  |  |  | Yes | Yes | Yes |  |  |  | Yes |  | Yes |  | Yes | Yes |  | 11 |
| Cigarette Smoking | Yes | Yes | Yes |  | Yes | Yes |  |  | Yes | Yes |  |  |  |  |  |  | Yes | Yes |  |  | Yes | Yes | 11 |
| Family History |  | Yes |  |  |  |  | Yes |  |  |  | Yes | Yes | Yes | Yes | Yes | Yes | Yes | Yes | Yes |  |  |  | 11 |
| Obesity |  | Yes | Yes | Yes | Yes |  | Yes | Yes |  | Yes |  |  |  |  |  |  | Yes |  |  |  |  | Yes | 9 |
| Sex | Yes | Yes |  |  | Yes |  |  |  |  |  | Yes | Yes | Yes |  |  |  |  |  |  |  | Yes | Yes | 8 |
| Blood Pressure | Yes | Yes | Yes | Yes | Yes | Yes |  |  |  | Yes |  |  |  |  |  |  |  |  |  | Yes |  |  | 8 |
| Alcohol | Yes | Yes | Yes | Yes |  |  |  |  |  | Yes |  |  |  |  |  |  |  |  |  |  | Yes |  | 6 |
| Medication exposure |  |  |  | Yes |  |  | Yes |  |  |  | Yes |  |  |  |  |  | Yes |  |  | Yes | Yes |  | 6 |
| Exposure to inhaled particulates |  |  |  |  |  |  |  | Yes | Yes | Yes | Yes |  |  |  |  |  | Yes | Yes |  |  |  |  | 6 |
| Social deprivation |  | Yes |  |  | Yes |  |  | Yes |  |  |  |  | Yes |  |  |  |  | Yes |  |  |  |  | 5 |
| Physical inactivity | Yes | Yes |  |  | Yes |  | Yes |  |  | Yes |  |  |  |  |  |  |  |  |  |  |  |  | 5 |
| Lower level of education. | Yes |  |  |  |  |  |  |  |  | Yes |  |  | Yes | Yes | Yes |  |  |  |  |  |  |  | 5 |
| Genetic or hereditary factors | Yes |  |  |  | Yes |  |  |  |  | Yes |  |  |  |  |  |  | Yes |  | Yes |  |  |  | 5 |
| Ethnicity |  | Yes |  |  | Yes |  | Yes |  |  |  |  |  |  |  |  | Yes |  |  | Yes |  |  |  | 5 |
| lack of social support |  | Yes |  |  |  |  |  |  |  | Yes |  | Yes | Yes |  |  |  |  |  |  |  |  |  | 4 |
| Cholesterol | Yes | Yes |  |  |  | Yes |  |  |  | Yes |  |  |  |  |  |  |  |  |  |  |  |  | 4 |
| Workplace exposures |  |  |  |  |  |  |  | Yes | Yes |  |  |  |  |  |  |  |  | Yes |  |  |  |  | 3 |
| Poor diet |  | Yes |  |  | Yes |  | Yes |  |  |  |  |  |  |  |  |  |  |  |  |  |  |  | 3 |
| Cannabis Use |  |  |  |  |  |  |  |  |  |  |  |  |  | Yes | Yes |  |  |  |  |  |  |  | 2 |
| Childhood adversity |  |  |  |  |  |  |  |  |  |  |  |  | Yes |  | Yes |  |  |  |  |  |  |  | 2 |
| Glomerular disease |  |  |  |  |  |  |  |  |  |  |  |  |  |  |  |  |  |  |  | Yes |  | Yes | 2 |
| Exposure (including prenatally) to tobacco smoke |  |  |  |  |  |  |  | Yes |  |  |  |  |  |  |  |  |  | Yes |  |  |  |  | 2 |
| Premature birth and associated low birth weight |  |  |  |  |  |  | Yes | Yes |  |  |  |  |  |  |  |  |  |  |  |  |  |  | 2 |

COPD: Chronic Obstructive Pulmonary Disease

PAD: Peripheral Arterial Disease

TIA: transient Ischaemic Attack
