## Supplementary material for "Identification of the Risk Factors for Complex-Multiple Long-term Conditions (C-MLTC): A Scoping Review": Suppl. Table 1b

| **Suppl. Table 1b**  **Risk factors for single long-term conditions (n=30)** | Stroke/TIA | Coronary Heart disease | Atrial Fibrillation | Heart Failure | Hypertension | PAD | Diabetes Mellitus | Asthma | COPD | Dementia | Parkinson’s Disease | Depression | Anxiety | Bipolar Disorder | Schizophrenia | Prostate Cancer | Ovarian Cancer | Lung Cancer | Breast Cancer | Chronic Kidney Disease (CKD) | Osteoporosis; | Rheumatoid Arthritis (RA). | **Total** |
| --- | --- | --- | --- | --- | --- | --- | --- | --- | --- | --- | --- | --- | --- | --- | --- | --- | --- | --- | --- | --- | --- | --- | --- |
| Development in-utero |  |  |  |  |  |  |  |  | Yes |  |  |  |  |  |  |  |  |  |  |  |  |  | 1 |
| Alpha-1-antitrypsin deficiency |  |  |  |  |  |  |  |  | Yes |  |  |  |  |  |  |  |  |  |  |  |  |  | 1 |
| Antiphospholipid syndrome and other hypercoagulable disorders. | Yes |  |  |  |  |  |  |  |  |  |  |  |  |  |  |  |  |  |  |  |  |  | 1 |
| Anticoagulation | Yes |  |  |  |  |  |  |  |  |  |  |  |  |  |  |  |  |  |  |  |  |  | 1 |
| Congenital heart disease | Yes |  |  |  |  |  |  |  |  |  |  |  |  |  |  |  |  |  |  |  |  |  | 1 |
| Influenza |  | Yes |  |  |  |  |  |  |  |  |  |  |  |  |  |  |  |  |  |  |  |  | 1 |
| Periodontitis |  | Yes |  |  |  |  |  |  |  |  |  |  |  |  |  |  |  |  |  |  |  |  | 1 |
| Excessive caffeine intake |  |  | Yes |  |  |  |  |  |  |  |  |  |  |  |  |  |  |  |  |  |  |  | 1 |
| Pregnancy |  |  |  | Yes |  |  |  |  |  |  |  |  |  |  |  |  |  |  |  |  |  |  | 1 |
| Anaemia |  |  |  | Yes |  |  |  |  |  |  |  |  |  |  |  |  |  |  |  |  |  |  | 1 |
| Thyrotoxicosis |  |  |  | Yes |  |  |  |  |  |  |  |  |  |  |  |  |  |  |  |  |  |  | 1 |
| Sepsis |  |  |  | Yes |  |  |  |  |  |  |  |  |  |  |  |  |  |  |  |  |  |  | 1 |
| Thiamine (Vit B1) deficiency |  |  |  | Yes |  |  |  |  |  |  |  |  |  |  |  |  |  |  |  |  |  |  | 1 |
| Salt |  |  |  |  | Yes |  |  |  |  |  |  |  |  |  |  |  |  |  |  |  |  |  | 1 |
| Gestational |  |  |  |  |  |  | Yes |  |  |  |  |  |  |  |  |  |  |  |  |  |  |  | 1 |
| Polycystic Ovary Syndrome |  |  |  |  |  |  | Yes |  |  |  |  |  |  |  |  |  |  |  |  |  |  |  | 1 |
| Metabolic Syndrome |  |  |  |  |  |  |  |  |  |  |  |  |  |  |  |  |  |  |  | Yes |  |  | 1 |
| Learning Disability |  |  |  |  |  |  |  |  |  | Yes |  |  |  |  |  |  |  |  |  |  |  |  | 1 |
| Hearing Impairment |  |  |  |  |  |  |  |  |  | Yes |  |  |  |  |  |  |  |  |  |  |  |  | 1 |
| Unemployed |  |  |  |  |  |  |  |  |  |  |  |  | Yes |  |  |  |  |  |  |  |  |  | 1 |
| Migration |  |  |  |  |  |  |  |  |  |  |  |  |  |  | Yes |  |  |  |  |  |  |  | 1 |
| Other cancers |  |  |  |  |  |  |  |  |  |  |  |  |  |  |  |  | Yes |  |  |  |  |  | 1 |
| Reproductive and hormonal factors |  |  |  |  |  |  |  |  |  |  |  |  |  |  |  |  | Yes |  |  |  |  |  | 1 |
| Ionising radiation |  |  |  |  |  |  |  |  |  |  |  |  |  |  |  |  |  | Yes |  |  |  |  | 1 |
| HIV/Aids |  |  |  |  |  |  |  |  |  |  |  |  |  |  |  |  |  |  |  | Yes |  |  | 1 |
| Acute Kidney Injury |  |  |  |  |  |  |  |  |  |  |  |  |  |  |  |  |  |  |  | Yes |  |  | 1 |
| Infections |  |  |  |  |  |  |  |  |  |  |  |  |  |  |  |  |  |  |  | Yes |  |  | 1 |
| Systemic lupus erythematosus (SLE) |  |  |  |  |  |  |  |  |  |  |  |  |  |  |  |  |  |  |  | Yes |  |  | 1 |
| Myeloma. |  |  |  |  |  |  |  |  |  |  |  |  |  |  |  |  |  |  |  | Yes |  |  | 1 |
| Gout |  |  |  |  |  |  |  |  |  |  |  |  |  |  |  |  |  |  |  | Yes |  |  | 1 |

COPD: Chronic Obstructive Pulmonary Disease

PAD: Peripheral Arterial Disease

TIA: transient Ischaemic Attack
