## Supplementary material for "Identification of the Risk Factors for Complex-Multiple Long-term Conditions (C-MLTC): A Scoping Review": Suppl. Table 2a

**Supplementary Table 2a. Systematic reviews of multimorbidity by condition.**

| **Study ID** | **Quality Assessment** | **Nos of studies** | **No. of participants** | **Types of participants** | **Outcomes reported** | **Multmorbidities reported** |
| --- | --- | --- | --- | --- | --- | --- |
| [Ayerbe 2017 [18]](https://pubmed.ncbi.nlm.nih.gov/28915505/) | Quality assessment tool of NIH | 5 | 7,626,964 | BPD or schizophrenia | HTN | BPD Schizophrenia |
| [Balomenakis 2023 [19]](https://pubmed.ncbi.nlm.nih.gov/37414144/) | CLARITY, GRADE | 17 | 3,149,547 | AF with cancer vs AF alone | Arterial thromboembolism | NR |
| [Barami 2017 [20]](https://pubmed.ncbi.nlm.nih.gov/28698089/) | NR | 12 | (unclear) | DM | GBM | NR |
| [Becker 2018 [21]](https://pubmed.ncbi.nlm.nih.gov/30339108/) | NOS & ROBINS-I | 10 | 38,650 | Anxiety | AD and vascular dementia | AD Vascular dementia |
| [Boutayeb 2013 [22]](https://pubmed.ncbi.nlm.nih.gov/23961989/) | NR | 26 | 104,182 | Patients with comorbidity or multimorbidity | Cancer, respiratory diseases, depression, mental disorders diabetes, CVD, MetS, HTN | Chronic respiratory disease, heart disease, hypertension, stroke, diabetes, rheumatism, peptic ulcer, kidney disease, hepatic disease and obesity |
| [Brain 2023 [23]](https://pubmed.ncbi.nlm.nih.gov/38455934/) | JBI | 25 MA | Unclear | CVD - HTN, AF, CCF, CHD | Dementia | Not specified |
| [Cai 2023 [24]](https://pubmed.ncbi.nlm.nih.gov/37728003/) | NOS, GRADE | 17 | 57,761 | Late-life depression or depressive symptoms | Stroke | NR |
| [Campbell 2021 [25]](https://pubmed.ncbi.nlm.nih.gov/33305479/) | NOS | 19 | 447,264 | DM | Hepatocellular CA | NR |
| [Chen 2015 [26]](https://pubmed.ncbi.nlm.nih.gov/26208998/) | QUADAS | 27 | 21,230,035 | COPD | HTN, DM, HF, CHD, PAD | NR |
| [Gaddam 2016 [27]](https://pubmed.ncbi.nlm.nih.gov/27881110/) | GRADE | 9 | 721,535 | COPD | CKD | NR |
| [Gan 2023 [28]](https://pubmed.ncbi.nlm.nih.gov/38074728/) | NOS | 18 | 29,694 | HTN | Depression | NR |
| [He 2024 [29]](https://pubmed.ncbi.nlm.nih.gov/38942197/) | QUIPS | 87 | 225,584 | At least 1 modifiable risk factor for MCI in community-dwelling adults aged 55 years or older | MCI | NR |
| [Hill 2021 [30]](https://pubmed.ncbi.nlm.nih.gov/34234373/) | Critical Appraisal Skills Programme | 37 | 595,911 | 2 or more of the following: Diabetes, heart disease, HTN, chronic lung disease, arthritis, heart failure, hyperlipidemia | Cognitive complaint | NR |
| [Hölzel 2011 [31]](https://pubmed.ncbi.nlm.nih.gov/20488546/) | Critical Appraisal Skills Programme | 25 | 5,192 | Anxiety disorder, personality disorder, HTN | Depression (chronic) | NR |
| [Huang 2020 [32]](https://pubmed.ncbi.nlm.nih.gov/33516963/) | Quality tool from the NHLB Institute | 61 | 11,810 | CKD and other risk factors | Anxiety | NR |
| [Huang 2020a [33]](https://pubmed.ncbi.nlm.nih.gov/33344074/) | Newcastle-Ottawa Scale (NOS) | 20 | 8,072,043 | COPD and another chronic condition | New-onset AF | COPD+DM: COPD+HTN: COPD+PVD: COPD+HF: COPD+CHF: COPD+Cancer: COPD+CKD: |
| [Huang 2024 [34]](https://pubmed.ncbi.nlm.nih.gov/39368643/) | STROBE-MR | 167 | Unclear | Multimorbidity, others | GI cancers | Not specified |
| [Ignacio 2024 [35]](https://pubmed.ncbi.nlm.nih.gov/38657829/) | AHRQ, NOS | 20 | 4,748 | Stroke | Depression, anxiety | Anxiety with depression |
| [Ikhile 2024 [36]](https://pubmed.ncbi.nlm.nih.gov/38551956/) | NOS | 21 | Unclear | Cancer | Depression, anxiety | See Comment |
| [Jamnitski 2013 [37]](https://pubmed.ncbi.nlm.nih.gov/22532629/) | NR (prevalence study) | 4 | 4,810 | Psoriatic arthritis | IHD, PVD, HF | NR |
| [Khaledi 2019 [38]](https://pubmed.ncbi.nlm.nih.gov/30903433/) | Unclear (not specified) | 248 | 83,020,812 | T2DM | Depression | NR |
| [Kuring 2020 [39]](https://pubmed.ncbi.nlm.nih.gov/32469813/) | Quality assessment tool of NIH | 120 | 127,274 | Clinically significant Depression, Anxiety, or PTSD | Dementia | Depression Anxiety PTSD |
| [Lanctôt 2023 [40]](https://pubmed.ncbi.nlm.nih.gov/38230722/) | None specified | 88 (SR + primary studies). 13 studies relevant for review | Unclear | AD dementia | DM, CVD, stroke | NR |
| [Lee 2023 [41]](https://pubmed.ncbi.nlm.nih.gov/37345504/) | JBI | 52 | 302,152 | Children, adolescent, and young adult patients with cancer (CYACs) | Depression, anxiety, psychotic disorders, suicide | See Comment |
| [Li 2020 [42]](https://pubmed.ncbi.nlm.nih.gov/32250298/) | QUADAS-2 | 12 | 16,200 | DM | Dementia in APOEɛ4 carriers | NR |
| [Meng 2024 [43]](https://pubmed.ncbi.nlm.nih.gov/39465033/) | NOS | 82 | 18,000,000 | COPD | IHD, Cardiovascular risk factors/comorbidities, cardiogenic shock, CHF, MACE | See Comment |
| [Mezuk 2008a [44]](https://pubmed.ncbi.nlm.nih.gov/19033418/) | NR | 7 | 119,685 | DM | Depression | NR |
| [Mezuk 2008b [45]](https://pubmed.ncbi.nlm.nih.gov/19033418/) | NR | 13 | 230,935 | Depression | DM | NR |
| [Mitchell 2013 [46]](https://pubmed.ncbi.nlm.nih.gov/22207632/) | PRISMA | 77 | 25,692 | Schizophrenia | MetS | NR |
| [Mourao 2016 [47]](https://pubmed.ncbi.nlm.nih.gov/26680599/) | NOS | 18 | 10,861 | Depression and MCI | Dementia | NR |
| [Naqvi 2019 [48]](https://pubmed.ncbi.nlm.nih.gov/30890833/) | NR | 29 | 1,704 | Rheumatoid arthritis | Depression | NR |
| [Naskar 2017 [49]](https://pubmed.ncbi.nlm.nih.gov/28558904/) | NR (prevalence study) | 41 | 34,119 | Indian patients with DM | Depression | NR |
| [Orlowski 2024 [50]](https://pubmed.ncbi.nlm.nih.gov/38653506/) | NR | 29 | Unclear | COPD | Multimorbidity | See Comment |
| [Osborne 2008 [51]](https://pubmed.ncbi.nlm.nih.gov/18817565/) | Unclear | 36 | 3,459,289 | Severe mental illnesses | DM, HTN, MetS | NR |
| [O'Sullivan 2021 [52]](https://pubmed.ncbi.nlm.nih.gov/33524598/) | NOS | 9 | 37,631 | T2DM, HTN, CKD | Colorectal CA | NR |
| [Ownby 2006 [53]](https://pubmed.ncbi.nlm.nih.gov/16651510/) | NOS | 20 | 102,172 | Depression | AD | NR |
| [Pahwa 2023 [54]](https://pubmed.ncbi.nlm.nih.gov/36538952/) | Modified published process (du Prel et al., 2009) | 7 | 875,302 | Bipolar disorder | T2DM | NR |
| [Pal 2018 [55]](https://pubmed.ncbi.nlm.nih.gov/30182156/) | Customized quality scoring tool | 12 | 6,865 | DM and MCI | Dementia | NR |
| [Peng 2020 [56]](https://pubmed.ncbi.nlm.nih.gov/32393205/) | Newcastle-Ottawa Scale (NOS) | 13 | 7,991,119 | COPD | T2DM | NR |
| [Polyakova 2014 [57]](https://pubmed.ncbi.nlm.nih.gov/24103852/) | Modified criteria published elsewhere (Luppa et al., 2012) | 23 | 23,083 | MCI | Depression (mild) | NR |
| [Romiti 2021 [58]](https://pubmed.ncbi.nlm.nih.gov/34333599/) | Customized tool based on the Newcastle–Ottawa Scale (NOS) | 46 | 4,200,000 | AF, COPD | HTN, DM, CHF, stroke/TIA, CAD | NR |
| [Roy 2012 [59]](https://pubmed.ncbi.nlm.nih.gov/23062861/) | NR (prevalence study) | 20 | Unclear: incl. primary studies and SRs | DM | Depression | NR |
| [Simayi 2019 [60]](https://pubmed.ncbi.nlm.nih.gov/31178523/) | AHRQ | 14 | 82,239,298 | T2DM | Depression | NR |
| [Steck 2018 [61]](https://pubmed.ncbi.nlm.nih.gov/29734098/) | Modified from Tooth et al., 2005 | 7 | 2,029 | AD and other chronic medical conditions | Depression | NR |
| [Tassew 2024 [62]](https://pubmed.ncbi.nlm.nih.gov/38917087/) | NOS | 12 | 4812 | HTN | Depression | NR |
| [Tazzeo 2023 [63]](https://pubmed.ncbi.nlm.nih.gov/37647994/) | NOS | 68 | Varied from 190 to 826,936 | Not specified: see comments | Multimorbidity | Not specified |
| [Tully 2014 [64]](https://pubmed.ncbi.nlm.nih.gov/25455809/) | Oxford Centre for Evidence-Based Medicine guidelines | 43 | 7,973 | CHD | Anxiety | Anxiety with depression |
| [Vrinzen 2023 [65]](https://pubmed.ncbi.nlm.nih.gov/36779863/) | Hoy's or O'Sullivan's | 161 | Unclear | Cancer | HTN, CHF, pulmonary disease, DM, dementia, renal disease, RA | NR |
| [Warner 2023 [66]](https://pubmed.ncbi.nlm.nih.gov/36709829/) | MMAT | 19 | 22,241 | Bipolar disorder | DM, cancer, renal disease, CVD, obesity | NR |
| [Xu 2013 [67]](https://pubmed.ncbi.nlm.nih.gov/23472134/) | NR | 15 | 8,496,332 | DM | Bladder cancer | NR |
| [Zafeiri 2021 [68]](https://pubmed.ncbi.nlm.nih.gov/30257266/) | GRADE | 24 | 205 | Diabetic peripheral neuropathy and depression | HTN, CHD | NR |

**Abbreviations**: AHRQ – Agency for Healthcare Research and Quality; BPD – bipolar depression; CHD – coronary heart disease; COPD – chronic obstructive pulmonary disease; GRADE – Grades of Recommendation, Assessment, Development and Evaluation; HTN – hypertension; MCI – mild cognitive impairment; MMAT – mixed methods appraisal tool; NOS – Newcastle Ottawa Scale.
