## Supplementary material for "Identification of the Risk Factors for Complex-Multiple Long-term Conditions (C-MLTC): A Scoping Review": Suppl. Table 2b

**Supplementary Table 2b. Systematic reviews quantifying risk of multimorbidity by condition.**

| **Study ID** | **Participants** | **Outcomes reported** | **Multmorbidities reported** | **Results: Risk of Long-term conditions: (odds ratio unless otherwise stated)** | | | | | | | | | | | |
| --- | --- | --- | --- | --- | --- | --- | --- | --- | --- | --- | --- | --- | --- | --- | --- |
|  |  |  |  | AF | HTN | DM | CHF | Stroke/TIA | CHD | PAD | Dementia | Depression | Anxiety | Cancer | CKD |
| [Ayerbe 2017 [18]](https://pubmed.ncbi.nlm.nih.gov/28915505/) | BPD or schizophrenia | HTN | BPD Schizophrenia |  | IRR 1.27 (1.15-1.40) IRR 0.94 (0.75 - 1.14) |  |  |  |  |  |  |  |  |  |  |
| [Balomenakis 2023 [19]](https://pubmed.ncbi.nlm.nih.gov/37414144/) | AF and cancer | Arterial thromboembolism |  |  |  |  |  |  |  | 0.97 (0.85-1.11) |  |  |  |  |  |
| [Becker 2018 [21]](https://pubmed.ncbi.nlm.nih.gov/30339108/) | Anxiety | AD and vascular dementia | AD Vascular dementia |  |  |  |  |  |  |  | HR 1.53 (1.16-2.01) OR 1.88 (1.05-3.36); |  |  |  |  |
| [Cai 2023 [24]](https://pubmed.ncbi.nlm.nih.gov/37728003/) | Late-life depression or depressive symptoms | Stroke |  |  |  |  |  | HR: 1.39 (1.22-1.58) |  |  |  |  |  |  |  |
| [Campbell 2021 [25]](https://pubmed.ncbi.nlm.nih.gov/33305479/) | DM | Hepatocellular CA |  |  |  |  |  |  |  |  |  |  |  | HR 1.26 (1.20-1.32) |  |
| [Chen 2015 [26]](https://pubmed.ncbi.nlm.nih.gov/26208998/) | COPD | HTN, DM, HF, CHD, PAD |  | 1.33 (1.13-1.56) | 1.36 (1.21-1.53) | 2.57 (1.90-3.47) |  | 1.86 (1.51-2.30) | 2.35 (1.48-3.74) |  |  |  |  |  |  |
| [Gaddam 2016 [27]](https://pubmed.ncbi.nlm.nih.gov/27881110/) | COPD | CKD |  |  |  |  |  |  |  |  |  |  |  |  | 2.20 (1.83-2.65) |
| [Huang 2020a [33]](https://pubmed.ncbi.nlm.nih.gov/33344074/) | COPD and another chronic condition | New-onset AF | COPD+DM: COPD+HTN: COPD+PVD: COPD+HF: COPD+CHF: COPD+Cancer: COPD+CKD: | 0.89 (0.54-1.49) 0.95 (0.70-1.28) 0.91 (0.89-0.94) 3.49 (2.70-4.52) 4.33 (2.47-7.59) 1.09 (0.92-1.29) 1.98 (1.74-2.25) |  |  |  |  |  |  |  |  |  |  |  |
| [Kuring 2020 [39]](https://pubmed.ncbi.nlm.nih.gov/32469813/) | Clinically significant Depression, Anxiety, or PTSD | Dementia | Depression Anxiety PTSD |  |  |  |  |  |  |  | 1.91 (1.72-2.12) 1.60 (1.29-2.00) 2.55 (0.43-15.12) |  |  |  |  |
| [Lee 2023 [41]](https://pubmed.ncbi.nlm.nih.gov/37345504/) | Children, adolescent, and young adult patients with cancer (CYACs) | Depression, anxiety |  |  |  |  |  |  |  |  |  | RR 1.57 (1.29-1.92) | RR 1.29 (1.14-1.47) |  |  |
| [Li 2020 [42]](https://pubmed.ncbi.nlm.nih.gov/32250298/) | DM | Dementia in APOEɛ4 carriers |  |  |  |  |  |  |  |  | RR 1.35 (1.13-1.63) |  |  |  |  |
| [Mezuk 2008a [44]](https://pubmed.ncbi.nlm.nih.gov/19033418/) | DM | Depression |  |  |  |  |  |  |  |  |  | RR 1.15 (1.02-1.30) |  |  |  |
| [Mezuk 2008b [45]](https://pubmed.ncbi.nlm.nih.gov/19033418/) | Depression | DM |  |  |  | RR 1.60 (1.37–1.88) |  |  |  |  |  | NR |  |  |  |
| [Mourao 2016 [47]](https://pubmed.ncbi.nlm.nih.gov/26680599/) | Depression and MCI | Dementia |  |  |  |  |  |  |  |  |  | RR 1.28 (1.09-1.52) |  |  |  |
| [Osborne 2008 [51]](https://pubmed.ncbi.nlm.nih.gov/18817565/) | Severe mental illnesses | DM, HTN, MetS |  |  | RR 1.11 (0.91 to 1.35) | RR 1.70 (1.21 to 2.37) |  |  |  |  |  |  |  |  |  |
| [Ownby 2006 [53]](https://pubmed.ncbi.nlm.nih.gov/16651510/) | Depression | AD |  |  |  |  |  |  |  |  | 2.02 (1.80–2.26) |  |  |  |  |
| [Pal 2018 [55]](https://pubmed.ncbi.nlm.nih.gov/30182156/) | DM and MCI | Dementia |  |  |  |  |  |  |  |  | 1.53 (1.20-1.97) |  |  |  |  |
| [Peng 2020 [56]](https://pubmed.ncbi.nlm.nih.gov/32393205/) | COPD | T2DM |  |  |  | RR 1.25 (1.16-1.34) |  |  |  |  |  |  |  |  |  |
| [Romiti 2021 [58]](https://pubmed.ncbi.nlm.nih.gov/34333599/) | AF, COPD | HTN, DM, CHF, stroke/TIA, CAD |  |  | 1.30 (0.97–1.73) | 1.80 (1.38–2.35) | 2.24 (1.73–2.90) | 1.18 (1.05–1.32) | 1.84 (1.44–2.35) |  |  |  |  |  |  |
| [Xu 2013 [67]](https://pubmed.ncbi.nlm.nih.gov/23472134/) | DM | Bladder cancer |  |  |  |  |  |  |  |  |  |  |  | RR 1.11 (1.00-1.23) |  |
