## Supplementary material for "Identification of the Risk Factors for Complex-Multiple Long-term Conditions (C-MLTC): A Scoping Review": Suppl. Table 3

**Supplementary Table 3. Long-term conditions as risk factors for Multimorbidity**

|  | **Target Condition** | | | | | | | | | | | | | | | | | |  |
| --- | --- | --- | --- | --- | --- | --- | --- | --- | --- | --- | --- | --- | --- | --- | --- | --- | --- | --- | --- |
| **Table 2:**  **Long-term conditions as risk factors for Multimorbidity** | Stroke/TIA | Coronary Heart disease | Atrial Fibrillation  (AF) | Heart Failure  (HF) | Hypertension | Peripheral Arterial Disease | Diabetes Mellitus | Asthma | Chronic Obstructive  Pulmonary Disease | Dementia | Parkinsons Disease | Depression | Anxiety | Bipolar Disorder &  Schizophrenia | Cancer | Chronic Kidney Disease | Osteoporosis; | Rheumatoid Arthritis (RA). | Total multimorbidity, |
|  | [**Link**](https://cks.nice.org.uk/topics/stroke-tia/background-information/risk-factors/) | [**Link**](https://cks.nice.org.uk/topics/cvd-risk-assessment-management/background-information/risk-factors-for-cvd/) | [**Link**](https://cks.nice.org.uk/topics/atrial-fibrillation/background-information/causes/) | [**Link**](https://cks.nice.org.uk/topics/heart-failure-chronic/background-information/causes/) | [**Link**](https://cks.nice.org.uk/topics/hypertension/background-information/risk-factors/) | [**Link**](https://cks.nice.org.uk/topics/peripheral-arterial-disease/background-information/risk-factors/) | [**Link**](https://cks.nice.org.uk/topics/diabetes-type-2/background-information/risk-factors/) | [**Link**](https://cks.nice.org.uk/topics/asthma/background-information/risk-factors/) | [**Link**](https://cks.nice.org.uk/topics/chronic-obstructive-pulmonary-disease/background-information/risk-factors/) | [**Link**](https://cks.nice.org.uk/topics/dementia/background-information/risk-factors/) | [**Link**](https://cks.nice.org.uk/topics/parkinsons-disease/) | [**Link**](https://cks.nice.org.uk/topics/depression/background-information/risk-factors/) | [**Link**](https://cks.nice.org.uk/topics/generalized-anxiety-disorder/background-information/risk-factors/) | [**Link**](https://cks.nice.org.uk/topics/bipolar-disorder/background-information/causes/) | [**Link**](https://cks.nice.org.uk/specialities/cancer/) | [**Link**](https://cks.nice.org.uk/topics/chronic-kidney-disease/background-information/causes/) | [**Link**](https://cks.nice.org.uk/topics/osteoporosis-prevention-of-fragility-fractures/background-information/risk-factors/) | [**0ink**](https://www.nice.org.uk/about/what-we-do/into-practice/measuring-the-use-of-nice-guidance/impact-of-our-guidance/nice-impact-arthritis/diagnosis-and-referral-of-inflammatory-arthritis) |  |
| **Risk factor** |  |  |  |  |  |  |  |  |  |  |  |  |  |  |  |  |  |  |  |
| Stroke/TIA | N/A |  |  |  |  |  |  |  |  | Y | Y | Y | Y |  |  | Y |  |  | 5 |
| Coronary heart disease | Y | N/A | Y | Y |  | Y |  |  |  | Y |  | Y | Y |  |  | Y |  |  | 8 |
| Atrial Fibrillation | Y | Y | N/A | Y |  | Y |  |  |  | Y |  |  | Y |  |  | Y |  |  | 7 |
| Heart Failure | Y |  | Y | N/A |  |  |  |  |  | Y |  | Y | Y |  |  | Y |  |  | 6 |
| Hypertension | Y | Y | Y | Y | N/A | Y |  |  |  | Y |  |  | Y |  |  | Y |  |  | 8 |
| Peripheral Arterial Disease | Y |  |  |  |  | N/A |  |  |  |  |  | Y | Y |  |  | Y |  |  | 4 |
| Diabetes Mellitus | Y | Y | Y |  | Y | Y |  |  |  | Y |  | Y | Y |  |  | Y | Y |  | 10 |
| Asthma |  |  |  |  |  |  |  |  | Y |  |  |  | Y |  |  |  |  |  | 1 |
| Chronic Obstructive Pulmonary Disease |  |  |  |  |  |  |  |  | N/A |  |  | Y | Y |  |  |  | Y |  | 3 |
| Dementia |  |  |  |  |  |  |  |  |  | N/A | Y | Y | Y |  |  |  |  |  | 3 |
| Parkinson’s Disease |  |  |  |  |  |  |  |  |  | Y | N/A | Y | Y |  |  |  |  |  | 3 |
| Depression |  |  |  |  |  |  |  |  |  | Y |  |  | Y |  |  |  |  |  | 2 |
| Anxiety |  |  |  |  | Y |  |  |  |  |  |  | Y | Y |  |  |  |  |  | 2 |
| Serious Mental Illnesses  (Bipolar Disorder and Schizophrenia) | Y | Y |  |  |  |  |  |  |  |  |  | Y | Y | N/A |  |  |  |  | 4 |
| Cancer  excluding non-melanoma skin cancers |  | Y | Y |  |  |  |  |  |  |  |  | Y | Y |  | Y | Y | Y |  | 7 |
| Chronic Kidney Disease (CKD) | Y | Y |  | Y | Y | Y |  |  |  |  |  | Y | Y |  | Y | N/A |  |  | 8 |
| Osteoporosis |  |  |  |  |  |  |  |  |  |  |  | Y | Y |  |  |  | N/A |  | 2 |
| Rheumatoid Arthritis (RA) | Y | Y |  |  |  |  |  |  |  |  |  | Y | Y |  |  |  | Y |  | 5 |
| Total number of conditions contributing to the risk of the target condition | 9 | 7 | 5 | 4 | 3 | 5 | 0 | 0 | 1 | 8 | 2 | 14 | 17 | 0 | 2 | 8 | 4 | 0 |  |
